# SARS-CoV-2 in environmental samples of quarantined households

**DOI:** 10.1101/2020.05.28.20114041

**Authors:** Manuel Döhla, Gero Wilbring, Bianca Schulte, Beate Mareike Kümmerer, Christin Diegmann, Esther Sib, Enrico Richter, Alexandra Haag, Steffen Engelhart, Anna Maria Eis-Hübinger, Martin Exner, Hendrik Streeck, Ricarda Maria Schmithausen

## Abstract

The role of environmental transmission of SARS-CoV-2 remains unclear. Particularly the close contact of persons living together or cohabitating in domestic quarantine could result in high risk for exposure to the virus within the households. Therefore, the aim of this study was to investigate the whereabouts of the virus and whether useful precautions to prevent the dissemination can be given.

21 households under quarantine conditions were randomly selected for this study. All persons living in each household were recorded in terms of age, sex and time of household quarantine. Throat swabs for analysis were obtained from all adult individuals and most of the children. Air, wastewater samples and surface swabs (commodities) were obtained and analysed by RT-PCR. Positive swabs were cultivated to analyse for viral infectivity.

26 of all 43 tested adults (60.47 %) tested positive by RT-PCR. All 15 air samples were PCR-negative. 10 of 66 wastewater samples were positive for SARS-CoV-2 (15.15 %) as well as 4 of 119 object samples (3.36 %). No statistically significant correlation between PCR-positive environmental samples and the extent of infection spread inside the household could be observed. No infectious virus could be isolated under cell culture conditions.

As we cannot rule out transmission through surfaces, hygienic behavioural measures are important in the households of SARS-CoV-2 infected individuals to avoid potential transmission through surfaces. The role of the domestic environment, in particular the wastewater load in washbasins and showers, in the transmission of SARS CoV-2 should be further clarified.

**Highlights:**

- With public “shut downs” due to SARS-CoV-2, domestic infection is a main possible route of transmission.
- All analysed air samples were tested negative for SARS-CoV-2.
- 15.15 % of all wastewater samples (washbasin, showers and toilets) were tested positive.
- Only 3.36 % of all object samples were tested positive: one remote control, two metallic door knobs and one wooden stove overlay.
- This study supports the hypothesis that indirect environmental transmission may only play a minor role, which needs clarifications in further studies.

## Introduction

The COVID-19 pandemic is one of the most important public health threats to the world since the Spanish flu around 100 years ago. Over 5 million cases and almost 350,000 deaths have been reported so far (WHO, 2020a, data as of 23rd may 2020). Thus, the COVID-19 pandemic challenges the environmental hygiene: special isolation and infectious disease wards have been established in hospitals and healthcare facilities and whole households have been quarantined. Hence, comprehensive monitoring of the environment of healthcare facilities and households during pandemic outbreaks are vital parts to ensure patients’ safety and public health (Liu et al., 2020).

COVID-19 is a disease of the upper airways (Schmithausen et al., 2020). It has been shown for SARS-CoV-2 that droplets (particles > 5 μm) can deposit on mucous surfaces of the upper respiratory tract and be spread when coughing, sneezing or speaking (Anfinrud et al., 2020; Li et al., 2020; Liu et al., 2020) Thus, the main airborne transmission pathway of infectious SARS-CoV-2 is aerosol (particles <5 μm) or droplets (van Doremalen et al., 2020). This is particularly important regarding indoor environments, because small particles with a higher viral load may be carried over distances up to 10 m from the emission source and may even accumulate (Morawska and Cao, 2020; Paules et al., 2020). However, these findings are based on laboratory experiments. One of a few on-field outbreaks studies on environmental transmission dynamics of SARS-CoV-2 was performed by Xu et al. (2020) on a cruise ship with a total of 3,711 passengers. Xu et al. (2020) rebutted that long-range airborne transmission routes and even central air conditioning systems play a role in a COVID-19 outbreak in a confined space. On the other hand it can be assumed that close contact and fomites contribute to transmission effects (ECDC, 2020; WHO, 2020a). Chan et al. (2020) proofed person-to-person transmission in hospital and family settings. Currently, only few on-field studies of SARS-CoV-2 detected RNA on door handles and surfaces in hospital and/or confirmed COVID-19 in the patient’s environment, particularly in Asia (Liu et al., 2020). However, the prevalence and potential transmission risks of SARS-CoV-2 in the environment of infected persons of the general population living with their families in households have not yet been sufficiently explored.

One important underlying question is whether and how long virus particles can survive on various surfaces to enable human-to-surface-to-human transmission. To date, no case of transmission of SARS-CoV-2 from human to human via food, drinking water or fomites could be demonstrated, although if there is speculation that in the early phase of virus spread in China transmission might be food-borne associated (Jalava, 2020). Modelling also implies that the indirect transmission of SARS-CoV-2 from environment is of little importance (Ferretti et al., 2020). In contrast, recent studies suggested that the environmental stability of the virus on surfaces plays an important role in the transfer (Otter et al., 2016; van Doremalen et al., 2020). Studies also showed that the viable virus remained detectable for hours or even days in the inanimate surroundings like the air, on stainless steel and plastic surfaces (Kampf et al., 2020; Service, 2020) as well as in urine and faeces of formerly positive patients (Holshue et al., 2020; Wang et al., 2020c; Wang et al., 2020b). SARS-CoV-2 RNA has been detected in the stool of one of the first patients in the USA (Holshue et al., 2020). This might be in line with the observation of Wang et al. (2020) and Cheng et al. (2020) describing that at least 2-10% of patients with COVID-19 show gastrointestinal symptoms such as diarrhoea and vomiting (Chen et al., 2020; Wang et al., 2020a). Schmithausen et al. (2020) described persistent diarrhoea in 32% of tested persons. Following the wastewater pathway, SARS-CoV-2 RNA has already been found in the wastewater of hospitals treating COVID-19 patients (Wang et al., 2020c). In a lab-based experiment, coronavirus (SARS-CoV-1) was found to remain infectious for 14 days at 4°C, and for 2 days at 20°C in hospital wastewater (Wang et al., 2005). Yeo et al. (2020) highlight the potential of faecal-oral transmission. Thus, wastewater and sanitation units represent potential infectious sources of SARS-CoV-2 and colonization of the sewage system with microorganisms already starts in the siphons of the washbasins, shower siphons, as well as in the toilets (KRINKO, 2020; Sib et al., 2019). When SARS-CoV-2 infected persons with gastrointestinal symptoms excrete urine and faeces, consequently SARS-CoV-2 can be identified in the immediate surroundings like the sanitary facilities. While the elimination of SARS-CoV-2 in wastewaters is possible after treatment (Holshue et al., 2020; Zhang et al., 2020), the possibility of faecal-oral recirculation of SARS-CoV-2 from siphons of washbasins and showers as well toilets to humans via droplets or aerosols or even smear-infection is still unclear. In order to test this assumption, the study presented here also collected on-field samples of siphons and toilets in private households of COVID-19 infected people.

The main exposure to and transmission of the virus occurs at home (Qian et al., 2020), and in cases of mild COVID-19 progression, home care is implemented to avoid hospital overload (WHO, 2020b). Consequently, contact persons of positively tested people are also placed in pre-emptive home isolation before the onset of symptoms due to the risk of contagion (ECDC, 2020; WHO, 2020b). As a result, infected people and contact persons living together as a family or in cohabitation are in domestic quarantine with each other. Even with separate bathrooms and bedrooms, it is impossible to effectively and permanently distance oneself and maintain adequate hand hygiene.

The aim of this study was to investigate the dissemination of virus in air, wastewater and on items within the domestic environment of family households with at least one SARS-CoV-2 positive family member, during a quarantine ordered by the local health department and to give useful recommendations for infection prevention.

## Material and Methods

### Sample site and recruitment of households

Samples were obtained in a high-prevalence community setting with Germany’s first largest high-prevalence cluster with regard to COVID-19 known at that point of time in March 2020 (Streeck et al., 2020b; Streeck et al., 2020a). The local health department provided lists of all positively tested inhabitants who had been placed in domestic quarantine at the point of data collection. 21 households, with at least one person tested positive for SARS-CoV-2 RNA, were randomly selected from this list. The respective residents were contacted by telephone and informed about the study. All persons and their family members living under one roof agreed to participate in the study. Complete information from pharyngeal swabs was available for all 58 study participants (43 adults, 15 children) living in 21 households.

### Sampling

Age, sex and time of quarantine were recorded for all individuals living in each household. Household was defined as people living together within one flat or one house and having regularly and close contact nearly every day. Throat swabs for virologic diagnostics were obtained from all adults described by Streeck et al. (2020b).

As this is an exploratory study, no standardised environmental sampling was carried out. Furthermore, no characterization of cleaning methods or materials was performed. Critical rooms and fomites were identified in each household by two researchers (physicians or virologists or hygienists or public health specialists) in cooperation with the residents. The focus of this study was on the air, wastewater and swab samples of as many different fomites (consumer goods and furnishings) as possible with the WHO “how to” guide as a reference (WHO, 2020c).

Air samples were obtained employing cyclone sampling (Verreault et al., 2008) via Coriolis Micro - Air sampler (Bertin Technologies SAS, France). The air collectors were positioned in the middle of the room that was used most frequently by the residents; this was usually the living room or the kitchen - all the rooms had no ventilation equipment. During sampling, close contact to the air sampler (e.g. speaking in a range below 2 metres but not above 3 metres) was avoided. Sampling was performed with 300 litres per minute for 10 minutes in 15 ml of 0.9 % NaCl.

Wastewater samples were obtained using sterile syringes and catheters to reach the wastewater in the siphons of sinks, showers and toilets in bathrooms. Samples were only taken when the sanitary facilities were shared between the residents. The air and wastewater samples were stored and transported at +4°C.

Fomite samples were taken using a swab with a synthetic tip and a plastic shaft (FLOQSwabs™, Copan, Italy) and added PBS, with 2 ml of 0.9 % NaCl including neutralizing buffer to counteract the effects of any residual disinfectant (WHO, 2020c). The residents identified fomites of frequent and shared use (e.g. door handles, remote control). All laboratory analyses were performed within 48 hours.

### Laboratory analysis

All samples were transported to the virologic laboratory within 6 hours of sampling. Virologic analysis was performed via RT-PCR using the protocol of Corman et al. (2020). Briefly, swab samples were homogenized by short vertexing and 140 μl of the sample were transferred to a sterile 2 ml microcentrifuge tube holding 560 μl AVL buffer (Qiagen). Viral RNA was extracted with the QIAamp Viral RNA Mini kit (Qiagen) according to the instructions of the manufacturer. The RNA was used as template for three real time RT-PCR reactions using SuperScript™ III One-Step RT-PCR System with Platinum™ TaqDNA Polymerase (Thermo Fisher) to amplify sequences of the SARS-CoV-2 E gene (primers E_Sarbeco_F and R, probe E_Sarbeco_P1), the RdRP gene (primers RdRP_SARSr_F, and R, and probe RdRP_SARSr-P2), and an internal control for RNA extraction, reverse transcription, and amplification (innuDETECT Internal Control RNA Assay, Analytik Jena #845-ID-0007100). Samples were considered positive for SARS-CoV-2 if amplification occurred in both virus-specific reactions.

The isolation of infectious virus from environmental samples was attempted by seeding Vero E6 cells in 24 well plates or T25 flasks at a density of 70-80 %. Cells were incubated with 200 μl (24 well) - 1000 μl (T25 flask) of the sample material supplemented with 1x penicillin/streptomycin/amphotericin B and incubated for 1 h at 37°C in 5 % CO_2_. For water samples, 10% (v/v) of inoculation volume was replaced by 10xPBS to obtain a final concentration of 1xPBS. After 1 h of incubation, the inoculum was removed, Dulbecco’s Modified Eagle’s medium (Gibco) with 3 % foetal bovine serum (Gibco) and 1x penicillin/streptomycin/amphotericin B was added. Cells were incubated over several days at 37°C, 5 % CO_2_ and observed for development of a cytopathic effect that typically occurs for growth of SARS-CoV-2 on Vero E6 cells.

### Statistical analysis

Statistical analysis was performed via Stata IC 15.1 (StataCorp, USA). An α = 0.05 was considered statistically significant and all tests were 2-tailed. The factors associated with environmental contamination were analysed using nonparametric tests for continuous variables. The χ^2^-Test or Fisher exact test were used to analyse categorical variables.

## Results

### Household data

In total, data from 21 households were included in the analysis. The profile of all investigated households is shown in table 1.

**Table 1:** Household data.

|  | Total | Per Household |  |  |
| --- | --- | --- | --- | --- |
|  |  | Median | IQR | Range |
| Number of households | 21 |  |  |  |
| Number of adults ($\geq 18$) | 43 | 2 | 2 – 2 | 1 – 4 |
| Number of children ($<18$) | 15 | 0 | 0 – 2 | 0 – 3 |
| Proportion of females (%) | 51.72 | 50.00 | 50.00 – 66.67 | 0.00 – 100.00 |
| Median household age (years) |  | 31.00 | 28.00 – 53.00 | 9.50 – 75.00 |
| Time of quarantine (days) |  | 5 | 5 – 6 | 0 – 6 |

Of the pharyngeal swab samples obtained from all 43 adults, 26 (60.47 %) tested positive by RT-PCR. The median number of adults testing positive was one per household (IQR: 1 - 2); in two households no PCR-positive person was discovered. We obtained samples from 9 children, with 4 of them tested positive (44.44 %). There was no association between positive adults and children within our study group (exact test, p = 0.469). The proportion of PCR- positive children was significant lower as the proportion of PCR-positive adults (binomial test, p = 0.016).

### Environmental sampling data

200 environmental samples (15 air samples [7.50 %], 66 wastewater samples [33.00 %], 119 object swabs [59.05 %]) from 21 households were included in the analysis. The median number of samples per household was 9 (IQR: 7 – 13, Min: 1, Max: 18). 14 samples (7.00 %) tested positive using RT-PCR. Overall, 14 samples (7.00 %) tested positive. Table 2 shows the number of PCR-positive samples considering the sample type. The observed differences in positivity between the sample types are significant (χ^2^-Test, p = 0.011). Infectious virus could not be isolated in Vero E6 cells from any environmental sample.

**Table 2:** PCR-status of different sample types.

| Sample type | PCR-negative | PCR-positive | Total number tested |
| --- | --- | --- | --- |
| Air samples | 15 (100 %) | 0 (0 %) | 15 (100 %) |
| Wastewater samples | 56 (84.85 %) | 10 (15.15 %) | 66 (100 %) |
| Object samples | 115 (96.64 %) | 4 (3.36 %) | 119 (100 %) |
| Total | 186 (93.00 %) | 14 (7.00 %) | 200 (100 %) |

As shown in table 2, wastewater samples were most commonly tested positive for SARS-CoV-2 RNA (15.15 %). For further analysis, four wastewater-subtypes were categorised: Washbasin siphons, shower siphons, toilet and process water. Table 3 shows the positive samples within these subtypes. No significance between wastewater subtype and detection of SARS-CoV-2-status was observed (χ_2_-Test, p = 0.700).

**Table 3:** PCR-status of wastewater sample subtypes.

| Sample subtype | PCR-negative | PCR-positive | Total number tested |
| --- | --- | --- | --- |
| Washbasin siphons | 21 (80.77 %) | 5 (19.23 %) | 26 (100 %) |
| Shower siphons | 13 (81.25 %) | 3 (18.75 %) | 16 (100 %) |
| Toilet | 21 (91.30 %) | 2 (8.70 %) | 23 (100 %) |
| Other | 1 (100 %) | 0 (0 %) | 1 (100 %) |
| Total wastewater samples: | 56 (84.85 %) | 10 (15.15 %) | 66 (100 %) |

In addition, the fomite samples were divided into six subtypes for further analysis: “Electronic devices”, “Knobs and handles”, “Plants and animals”, “Furniture and furnishings”, “Foods and drinks” and “Clothing”. Table 4 shows the results of PCR analysis within the subtypes. There was no significant association between object subtype and PCR-status (χ^2^-Test, p = 0.843).

**Table 4:** PCR-status of different fomite sample subtypes.

| Sample subtype | PCR-negative | PCR-positive | Total number tested |
| --- | --- | --- | --- |
| Electronic devices | 51 (98.08 %) | 1 (1.92 %) | 52 (100 %) |
| Knobs and handles | 29 (93.55 %) | 2 (6.45 %) | 31 (100 %) |
| Plants and animals | 11 (100 %) | 0 (0 %) | 11 (100 %) |
| Furniture and furnishing | 18 (94.74 %) | 1 (5.26 %) | 19 (100 %) |
| Foods and drinks | 4 (100 %) | 0 (0 %) | 4 (100 %) |
| Clothing | 2 (100 %) | 0 (0 %) | 2 (100 %) |
| Total object samples: | 115 (96.64 %) | 4 (3.36 %) | 119 (100 %) |

Four fomite samples tested positive (3.36 %), i.e. an electronic device (remote control), two metallic doorknobs and one wooden stove overlay.

No significant association between positive wastewater samples and positive object samples was observed (χ^2^-Test, p = 0.851, data not shown).

### Associations between human and environmental data

No statistically significant correlation could be observed between the household information collected and the detection of SARS-CoV-2 RNA in the environmental samples (χ^2^-Test, p = 0.148). The households with positive environmental PCR results were further analysed with regard to the number of adults (χ^2^-Test, p = 0.249), the number of children (χ^2^-Test, p = 0.263), the proportion of females (χ^2^-Test, p = 0.410), the median age per household (χ^2^-Test, p = 0.453) and the time of quarantine (χ^2^-Test, p = 0.459). No correlation between PCR-positive environmental samples and PCR-positive human samples could be found in this study (χ^2^- Test, p = 0.756). There was no household with PCR-positive environmental samples and PCR-negative human samples.

## Discussion

The results indicate that at that early time of SARS-CoV-2 outbreak research in Germany the contamination of the domestic environment is negligible during quarantine measured with the current state of the art methods. We could not detect any viral RNA in air samples and only 3.36 % of all fomite samples. In contrast, 15.15 % of all wastewater samples were positive for SARS-CoV-2 RNA, which indicates that mouthwash in washbasins, body wash in the shower and faeces in toilets and therefore wastewater could pose a relevant exposure (Wu et al., 2020). Although RT-PCR is a highly sensitive detection method it does not yield information on the infectivity of the virus in these samples. Attempts to isolate virus in cell culture were not successful. Given the rather low cycle threshold (CT) values >30 obtained in the RT-PCR analysis of these samples, the amount of potential virus is estimated to be too low for virus isolation in general. Indeed, virus isolation in cell culture has not been successful in our laboratory at a CT value >30 so far. Furthermore, several wastewater samples had a toxic effect on the cells, which might be linked to detergent residues. It is therefore difficult to give specific hygienic behaviour precautions but rather basic hygiene measures for dissemination prevention (KRINKO, 2020).

With regard to the fomite and surface samples, only few positive PCR results were found in this study. This might also be due to methodological problems. On the one hand, the swab/transport solution combinations used could not have been suitable for keeping viral RNA stable until it was analysed in the laboratory. Ideally, object swabs should cover 25 cm^2^ and be put into 2 ml of viral transport medium including neutralizing buffer to counteract the effects of any residual disinfectant or degrading enzymes (WHO, 2020c). On the other hand, we observed positive PCR results in the throat swabs that were collected by the same swab/transport combinations. Assuming that the results are not methodologically inappropriate, they could indicate that the environmental survival of SARS-CoV-2 may not be too long in the domestic environment. The survival times of SARS-CoV-2 on various dry materials for different periods of time (< 3 hours on printing and tissue papers, < 2 days on wood and clothing, < 4 days on smooth surfaces, < 7 days on steel or plastic) were investigated by (Chin et al., 2020). However, it should be noted that these data were generated under laboratory conditions. It can be assumed that households in quarantine have a cleaning regime, but even under these conditions viral RNA could be found on fomites in the households. A further characterization of different cleaning systems (frequency, cleaning agents, ventilation of rooms, etc.) would be necessary; however, this effect could only be validly estimated in observational studies, since a large bias towards social desirability can be expected in surveys.

Following international recommendations, air samples should be taken as swabs of ventilation exits or air purifier ventils (WHO, 2020c). Since this is just a surrogate for real air contamination and normally households in Germany are not equipped with ventilators or air purifiers, cyclone air samplers were used. Cyclone samplers may be less efficient than other sampler types at recovering low concentrations of airborne viruses due to the physical stress caused by centrifugal force (Bourgueil et al., 1992). However, a recent study using a cyclone air collector to investigate air contamination in isolation rooms of a hospital (Chia et al., 2020) showed that 2 out of 3 collected samples were positive for SARS-CoV-2 RNA. Further experimental investigations of different air samplers in defined environments and preferably concerning general population households would be necessary to exclude a method-related false low recovery rate. However, these findings suggest that droplet transmission is the main pathway of transmission and that aerosol transmission plays a rather minor role. According to this, droplets and/or aerosols with SARS-CoV-2 in a viable and infectious form can be formed while flushing open toilets without closed lids or arise from contaminated siphons and thus could become a transmission pathway.

With regard to the results of the wastewater samples in our study, the percentage of positive samples was lowest in toilets (8.70 %), higher in shower siphons (18.75 %) and highest in washbasin siphons (19.23 %). Although these differences were not found to be significant, they support the above-mentioned hypothesis that aerosolization of viral loaded droplets from these wastewater reservoirs can be possible. Even more, the viral load on the hands and in the throat is highest and viral particles can be released from spitting into the washing basin siphon after teeth brushing or hands washing. The excretion of SARS-CoV-2 RNA via faeces and urine has already been described (van Doremalen et al., 2020) and could also lead to an increased detection rate of viral RNA in the shower or toilet.

Thus, the wastewater system could serve as a possible surveillance system for the circulation of the virus within several environments (Medema et al., 2020). In general, wastewater requires special hygienic attention, for example with regard to multidrug-resistant bacteria and antibiotic residues (Müller et al., 2018; Sib et al., 2019; Voigt et al., 2020; Voigt et al., 2019), enteric viruses like norovirus or rotavirus (Lodder and Roda Husman, 2005) and coronavirus (Gundy et al., 2009). The enteric transmission of SARS-CoV led to a large outbreak cluster in Hongkong in 2003 (Leung et al., 2003). In addition, enteric dissemination of and exposure to SARS-CoV-2 via wastewater is also considered to be a main risk (Lodder and Roda Husman, 2020). Therefore, existing hygiene recommendations (washing hands after contact with wastewater, flushing the toilet with closed lid, avoiding re-contamination of drinking water systems and domestic environment by wastewater) are considered to be necessary to sufficiently control this transmission route. Furthermore, preventive and intervention measures should not start at the wastewater treatment in the treatment plant, but already in the immediate surroundings of the patient, in order to minimize the infection potential.

## Conclusions

The domestic environment predominantly does not seem to pose a high risk for transmission of SARS-CoV-2. Surfaces in the domestic environment did not show a high contamination rate in this study, whereas the detection of viral RNA in wastewater of washbasins, showers and toilets showed a significantly higher contamination with SARS-CoV-2, indicating a possible reservoir that has not been considered so far. However, further systematic studies with an adapted methodology should be performed to investigate the contamination of the domestic environment and the interactions between humans, animals and the environment. Furthermore, the possibility of transmission via wastewater has hygienic implications for systematic prevention measures especially for areas with poor sanitation (Carducci et al. 2020).

## Data Availability

not applicable

## Acknowledgements

We thank all persons and families for their active and committed participation in this study. We thank the team of Dr. H. Schößler of the Local Health Authority in Heinsberg for their cooperation and trust. We thank S. Pusch, the chief administrator of the Heinsberg district, and his supportive and professional team for giving us the opportunity to perform this study. We also thank Professor G. Bierbaum for giving expert advice. We gratefully acknowledge S. Tietjen and L. Sommer for on-site support. We are indebted to the Institute of Animal Sciences of the Agricultural Faculty of the University of Bonn for providing technical support.

